# High-Intensity Interval Training Outperforms Moderate Exercise to Improve Aerobic Capacity in Patients with Recent-Onset Idiopathic Inflammatory Myopathies: A Multicenter Randomized Controlled Trial

**DOI:** 10.1101/2024.12.05.24318130

**Authors:** Kristofer M. Andreasson, Cecilia Leijding, Maryam Dastmalchi, Antonella Notarnicola, Stefano Gastaldello, Takashi Yamada, Heléne Sandlund, Dag Leonard, Håkan Westerblad, Ingrid E. Lundberg, Daniel C. Andersson, Helene Alexanderson

**Affiliations:** Karolinska University Hospital, Theme Women’s Health and Allied Health Professionals, Medical Unit Allied Health Professionals, Stockholm, Sweden; Karolinska Institutet, Department of Medicine, Solna, Division of Rheumatology, Stockholm, Sweden; Karolinska Institutet, Department of Physiology and Pharmacology, Stockholm, Sweden; Karolinska University Hospital, Department of Gastro, Dermatology and Rheumatology, Theme Inflammation and Aging, Stockholm, Sweden; Sapporo Medical University, Graduate School of Health Sciences, Sapporo, Japan; Uppsala University, Department of Medical Sciences, Section of Rheumatology, Uppsala, Sweden; Karolinska University Hospital, Theme Heart, Vascular and Neuro, Cardiology unit, Stockholm, Sweden

## Abstract

**Objective:** To investigate efficacy, safety, and tolerance of high-intensity interval training (HIIT) versus standard low-moderate intensity home-based exercise (CON) to improve aerobic capacity, muscle endurance, and mitochondrial function in patients with recent onset, idiopathic inflammatory myopathies (IIM).

**Methods:** Twenty-three patients with recent onset IIM were randomized into two groups: HIIT and CON. HIIT underwent 12 weeks of supervised high-intensity training, while CON followed a low-moderate intensity home-based exercise program. Outcomes included maximal exercise test (VO_2_peak, peak power, time-to-exhaustion (TTE)), mitochondrial protein expression in muscle biopsy serum levels of muscle enzymes (CK, LD, AST, ALT), muscle strength (MMT8) and disease activity (subset of MDAAT), before and after intervention.

**Results:** HIIT resulted in a 16 % increase in VO_2_peak L/min, significantly higher than the 1.8 % change in CON (95 % CI 0.1;0.47). Peak power and TTE also improved significantly more in HIIT, 18 % and 23 %, respectively, compared to CON, 8 % and 12 %, (95 % CI 3.9;30.8 and 00:06;03:18, respectively). Muscle biopsies (HIIT n=7, CON n=6) showed increases (p<0.05) in central mitochondrial protein expression in HIIT but not in CON, suggesting enhanced mitochondrial function. Both groups maintained stable serum muscle enzymes indicating no increase in disease activity from the intervention. Muscle disease activity remained low and unchanged in both groups (95 % CI −1.2;1.0), physician global activity and MMT8significantly improved within CON (95 % CI −1.7;-0.26 and 0.1;3.9, respectively) but not in the HIIT group. Compliance with HIIT was high, and no adverse events were reported.

**Conclusion:** HIIT is a highly effective and safe exercise intervention to improve aerobic fitness, muscle endurance, and mitochondrial function in patients with recent onset IIM. This approach should be considered as an adjuvant treatment in managing IIM, potentially enhancing the health for these patients.

**The study was registered at ClinicalTrials.gov: NCT03324152**

**Significance and Innovations:**

1. High-intensity interval training (HIIT) is more effective to improve aerobic capacity (peak oxygen uptake; VO_2_peak), increasing 16 % compared to 1.8 % change in the moderate exercise control group in patients with recent onset IIM.
2. HIIT produced larger exercise adaptations with increased expression of aerobic metabolism (mitochondrial) proteins in muscle tissue compared to moderate exercise.
3. HIIT is a safe and well tolerated exercise intervention for patients with recent onset IIM.

## Introduction

Idiopathic inflammatory myopathies (IIM) are a heterogenous group of autoimmune disorders typically characterized by chronic muscle inflammation. Muscle weakness and reduced muscle endurance as well as systemic features including inflammation in extra-muscular organs such as lungs, skin, heart, and joints are common. In addition, pain and fatigue are frequently reported (1). Medical treatment consists of initially high dose of oral glucocorticoids in combination with steroid sparing agents, such as methotrexate with add-ons depending on treatment response, organ involvement and disease severity. The treatment response and prognosis vary, and although relying on pharmacological treatment, exercise is an adjuvant treatment and imperative to effectively manage IIM (2, 3).

Patients with IIMs typically suffer from muscle weakness, fatigue, and reduced endurance capacity. At diagnosis, patients have approximately 70 % reduced muscle endurance compared to reference values as measured by the Functional Index-2 (FI-2), and 6 % reduced maximal strength as measured by the Manual Muscle Test 80 (MMT8) (4). Whether being due to maximal muscle strength or muscle endurance, the impaired muscle function is often sustained and rarely recovers to pre-disease levels leading to permanent limitations in daily life.

The cause of muscle impairment is less understood but is thought to involve immune and non-immune mechanisms. Although inflammatory cells are often seen in muscle tissue, the level of inflammation in the muscle does not correlate with reduced muscle strength or impaired endurance (5, 6). Several mechanisms have been proposed and point to a multifactorial model that includes tissue remodeling (e.g. capillary dysfunction (7) and fibrosis (8)), immune mediated mechanisms (9, 10), and impaired myocyte functions (e.g. contractility, mitochondrial dysfunction and endoplasmic reticulum (ER) stress (11, 12, 13)). Impaired mitochondrial function might be indicated by the observation of cytochrome C oxidase (COX) negative fibers often observed in muscle biopsies from patients with IIM. Impaired mitochondrial function will be a factor of reduced aerobic capacity as the ability to use oxygen for energy production in the muscle will be limited (14). Further, the capillary density can be reduced, specifically in patients with DM, which may reduce oxygen supply to the muscle tissue (15, 16). Physical inactivity and side effects of drug treatment may also contribute to muscle weakness (17, 18).

Maximal oxygen consumption (VO_2_peak) is a critical marker for aerobic fitness that correlates to overall survival and muscle aerobic metabolism (19, 20). The VO_2_peak depends on cardiac output and the peripheral arterio-venous oxygen extraction that depends on the micro circulation and metabolic function in the working muscle (e.g., mitochondrial oxygen extraction). There is a close relationship between VO_2_peak and muscle mitochondrial function, and VO_2_peak can effectively be increased with exercise training including high-intensity interval training (HIIT) in healthy individuals (19, 20). In patients with IIM (i.e., PM, DM), VO_2_peak is known to be reduced by approximately 25-50 % (6, 21). The mechanism of reduced VO_2_peak in IIM is not clear. Yet, it is independent of the presence of lung diseases in these patients (22, 23) and likely depend on reduced peripheral O_2_ extraction and/or cardiac output.

### Exercise studies in IIM

Exercise was once considered harmful for patients with IIM due to an imagined risk of aggravated myofiber damage. However, this concept has been rebutted by numerous studies that show safety and clinical benefits of mild-to moderate exercise (24). Further, moderately intensive endurance exercise can reduce disease activity and inflammation as well as improve mitochondrial enzyme activity in patients with longstanding inflammatory low-active IIM (21, 25). Out of the exercise studies published, randomized controlled trial designs are rare (24). The only RCT on exercise conducted in patients with recent onset IIM, showed that easy to moderate intensity home exercise in combination with medical treatment is safe but did not outperform medical treatment alone in the non-exercising control group, likely due to the use of low-moderate intensity training protocol (26, 27). However, the confirmation of safety of the home exercise program at time of diagnosis has led to the use of this training protocol as standard of care adjunct therapy for IIM patients in Sweden. However, it is unclear if exercise training on high intensity levels could effectively, and safely, improve physical capacity in patients with recent onset IIM.

## Hypotheses and objectives

Our hypothesis is that exercise on high intensity is an effective intervention to improve aerobic fitness and induce muscle adaptations in IIM. The previously evaluated exercise protocols in IIM have been of a too low intensity level to induce adequate physiological adaptations. Moreover, we believe that protocols with continuous and longer exercise sessions, although on lower intensity may be less feasible in recent-onset IIM due to substantial fatiguability seen in this patient group. In contrast, high intensity short interval-based exercise may be more tolerable, render more muscle adaptations and thereby being more effective to improve exercise capacity in recent-onset IIM patients. We hypothesized that HIIT would be tolerated for patients and lead to increased VO_2_peak. Secondary, we hypothesized that HIIT would induce beneficial adaptations to muscle metabolism in IIM.

This multicenter randomized controlled trial compared 12 weeks of HIIT to the current standard of care low-moderate intensity home-based exercise. The primary objective was to test the hypothesis that HIIT is more effective to improve aerobic capacity in patients with recent onset IIM, and that it still meets safety standards. In conjunction to this we tested the hypothesis that HIIT may lead to aerobic adaptations in the IIM muscle with increase in mitochondrial proteins.

## Materials and methods

This study was approved by the Regional Ethics Review Board in Stockholm (Dnr: 2016/2444-31), who also approved two amendments regarding serum sample and including Akademiska Hospital in Uppsala (Dnr: 2018/1350-32 and 2018/2614-32. The project is registered at ClinicalTrials.gov (ID NCT03324152). All participating patients signed an informed consent before being included.

### Patients

All newly diagnosed patients with any type of adult IIM, except inclusion body myositis (IBM), registered at the Karolinska University Hospital in Stockholm between October 2017 – April 2023, and at Akademiska Hospital in Uppsala between February 2019 – April 2023, who fulfilled the inclusion criteria, were invited to participate.

The inclusion criteria were probable or definite PM or DM according EULAR/ACR 2017 Criteria for IIM (28), antisynthetase syndrome (ASyS) according to Connors Criteria (29), or immune-mediated necrotizing myopathy (IMNM) according to 2016 ENMC Criteria (30), 18-70 years old, < 12 months of diagnosis duration and medical treatment.

Exclusion criteria were: IBM according to the 2013 ENMC criteria (31), active cancer, heart- or lung involvement or disease contraindicating intensive exercise (e.g., severe interstitial lung disease, oxygen supplementation or myocarditis), severe muscle weakness, pain or general fatigue hindering participation in HIIT, and osteoporosis.

A power analysis of the first six participants’ VO_2_peak revealed a necessity >21 participants to reach statistical power (β = 0.2 and α = 0.05). Twenty-three patients, out of 82 screened, were enrolled. Out of the 59 not included, 24 did not meet inclusion criteria, or met exclusion criteria, 15 declined participation and 20 were not included due to other reasons (e.g., not responding calls, not speaking Swedish or English, living in a different region even though diagnosed in our clinic), see recruitment flow chart (Figure 1).

**Figure 1.**
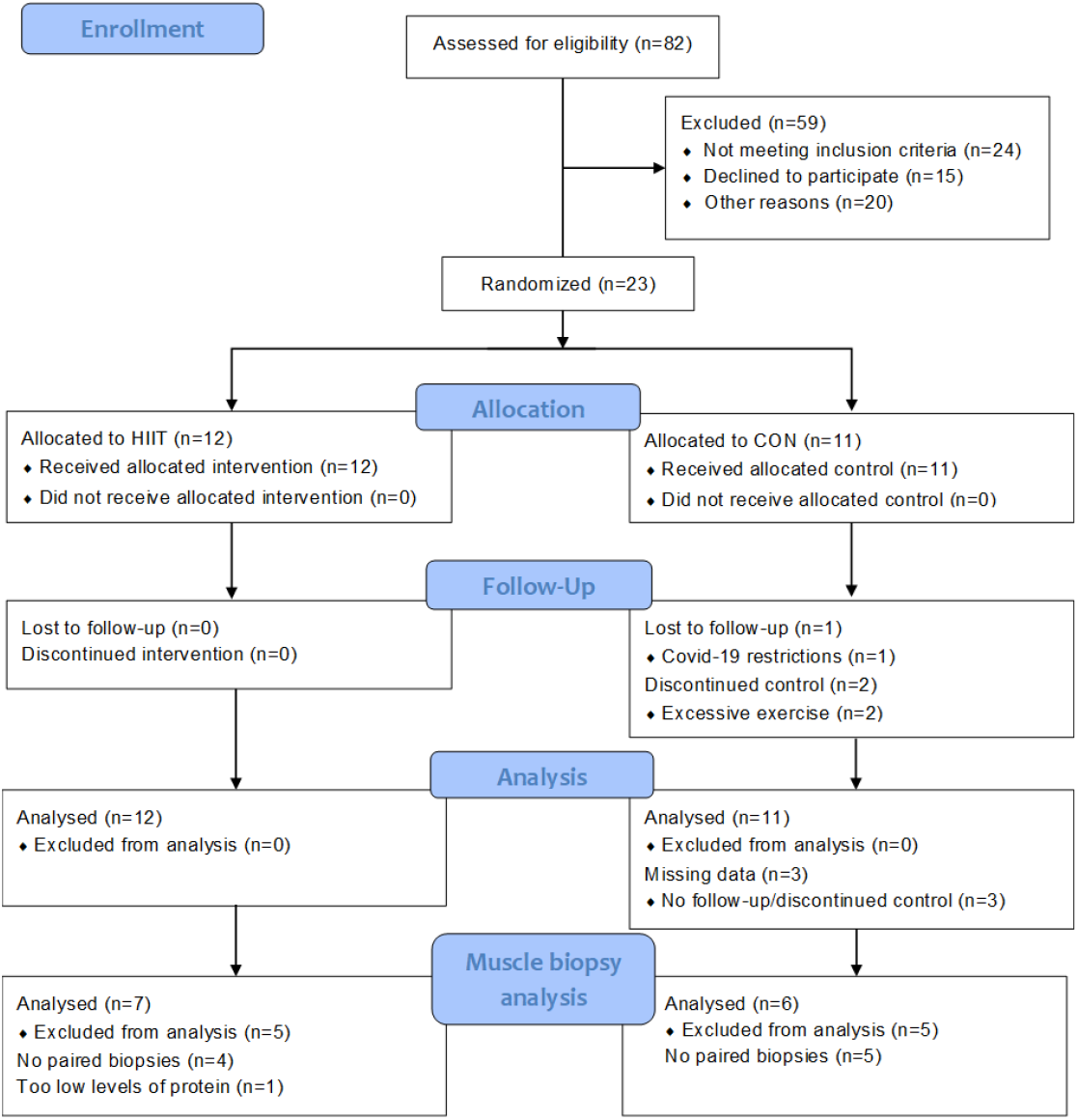
Recruitment flowchart. HIIT, high-intensity interval training; CON, control group.

If there was an uncertainty of the patient’s ability to perform the intensive exercise program, discussion within the medical team consisting of medical doctors (rheumatologists and cardiologist, and other specialties if needed), patient responsible doctor and physical therapists. For baseline characteristics see Table 1.

**Table 1.** Demographics of 23 patients before and after 12 weeks of exercise.

| Variables | HIIT |  | CON |  |
| --- | --- | --- | --- | --- |
|  | Baseline<br>n=12 | 12-week follow-up<br>n=12 | Baseline<br>n=11 | 12-week follow-up<br>n=8 |
| Sex F/M, n | 6/6 | 6/6 | 9/2 | 7/1 |
| Diagnosis PM/DM/ASyS, n | 4/4/4 | 4/4/4 | 0/6/5 | 0/4/4 |
| Age, years (min-max) | 34.5 (24–65) |  | 57 (41–64) |  |
| Weight, kg (IQR) | 77 (71.5–83) | 78 (72–82) | 67 (58–82) | 66 (63–76) |
| Disease duration, months (min-max) | 5 (3–11) |  | 5 (1–8) |  |
| Symptom to diagnosis, months (min-max) | 10 (1–44) |  | 3.5 (1–17) |  |
| Pulmonary disease activity > 0 cm, n* | 6 | 3<sup>1</sup> | 4<sup>1</sup> | 2 |
| Pulmonary disease activity, 0–10 (min-max) | 0.45 (0.0–2.5) | 0 (0.0–1.8)<sup>1</sup> | 0 (0–2.5)<sup>1</sup> | 0 (0–3.1)<sup>3</sup> |
| DMARD AZA/MMF/HYD/MTX/SEND/RIT (n total) | 1/5/X/4/X/6 (16) | 1/6/X/5/X/5 (17) | X/4/1/3/1/3 (12) | X/4/2/3/X/2 (10) |
| Prednisone, mg/day (IQR) | 10.6 (4.7–22.5) | 5 (0–9) | 15 (12.5–17.5)<sup>2</sup> | 6.2 (3.5–10.5) |
| CK, μkat/L (min-max) | 1.5 (0.6–10.2) | 1.5 (0.5–11.3) | 1.2 (0.42–2.2)<sup>2</sup> | 1.5 (0.42–2.8)<sup>1</sup> |
| LD, μkat/L (min-max) | 3.7 (2.9–6.9) | 3.7 (2.4–6.6) | 3.7 (3.1–6)<sup>2</sup> | 3.7 (2.7–6)<sup>1</sup> |
| ASAT, μkat/L (min-max) | 0.42 (0.26–1.44) | 0.41 (0.25–1.97) | 0.34 (0.26–1.47)<sup>2</sup> | 0.42 (0.32–0.55)<sup>1</sup> |
| ALAT, μkat/L (min-max) | 0.47 (0.13–0.98) | 0.40 (0.21–1.23) | 0.41 (0.14–0.92)<sup>2</sup> | 0.31 (0.25–1.37)<sup>1</sup> |

Data presented in median except when absolute numbers as specified by n. HIIT, high-intensity interval training; CON, control; F, female; M, male; PM, polymyositis; DM, dermatomyositis; ASyS, antisynthetase syndrome; DMARDs, disease modifying anti-rheumatic drugs; AZA, azathioprine; MMF, mycophenolate mofetil; HYD, hydroxychloroquine; MTX, methotrexate; SEND, sendoxan; RIT, rituximab; mg, milligram; CK, creatine phosphokinase; $\mu$ kat/L, microkats per liter; LD, lactate dehydrogenase; ASAT, aspartate transaminase; ALAT, alanine transaminase. Normative values male/female: CK 0.8–6.7/0.6–3.5, LD <3.5/<3.5, ASAT <0.76/<0.61, ALAT <1.1/<0.76. 1 $\mu$ kat/L is equal to 60 IU/L. Superscript numbers indicate n of missing data. \*Types of pulmonary diseases: antisynthetase autoantibody associated interstitial lung disease (anti-ARS-ILD), myositis-related nonspecific interstitial pneumonia (NSIP), diffuse alveolar hemorrhage, hilar enlargement, ground glass changes, fibrosis.

## Assessments

### Clinical visit

At enrollment the patient visited their treating rheumatologist and the myositis team nurse. Blood samples for serum levels of muscle enzymes (creatine kinase (CK), lactate dehydrogenase (LD), aspartate aminotransferases (AST) and alanine aminotransferases (ALT)) as surrogate markers for inflammation were taken according to clinical routine, Manual Muscle Test 80 (MMT8), physician global assessment (PhGA) and MDAAT muscle disease activity were performed, to investigate tolerance.

### Fitness tests

#### Maximal exercise test

VO_2_peak L/min was the primary outcome and was assessed by a maximal exercise test performed on a stationary bike using a standardized protocol for all participants. VO_2_peak was measured breath-by-breath gas exchange with Innocor model INN00500 (COSMED) (32). First, a resting electrocardiogram (ECG) was registered, then biking started on 30 W with a step increase of 10 W per minute until exhaustion. The participant was instructed to keep cycling until exhaustion, and when unable to keep 60-65 RPM the test was stopped. Staff could stop the test if appropriate due to live-ECG or participant-reported symptoms (e.g., dizziness or chest pain). Every minute blood pressure was measured, and the participant rated the perceived exertion (Borg’s RPE, 6-20), breathlessness and chest pain (Borg’s CR10, 0-10). The test was followed by a recovery period of a minimum of six minutes. The primary outcome of this test was VO_2_peak in liters/minute (L/min) and secondary outcomes were peak load in watts (peak power), peak heart rate (HRpeak), and time-to-exhaustion (TTE), which was the time measured from start to finish of the test (peak power and TTE being measures of muscle endurance). The test was performed approximately one week after the last exercise session and was led by independent staff at the physiology department who were unaware of group allocation; two biomedical scientists, one handling the Innocor assessment and one in charge of the rest of the test. A cardiologist was always present or immediately available during the test.

### Muscle biopsy and analysis

#### Biopsy procedure

Muscle biopsies were taken at diagnosis (PRE) and as follow-up post exercise intervention (POST) using the percutaneous conchotome technique under local anesthesia in the outpatient clinic (33). A 5–10-millimeter incision is done and then two to four samples of muscle tissue, the size of 3×8 mm maximum, are taken. Biopsy sites were vastus lateralis or tibialis anterior depending on the participant’s conditions (e.g., amount of fat tissue or antithrombotic treatment), and at contra-lateral side of the same muscle at follow-up (within one week of the final session). The muscle piece was frozen immediately by dipping it directly into the freezing medium. The piece was placed in a tube with a few frozen drops of plain water to hinder it from drying and stored at −80°C. The freezing medium was initially isopenthane chilled by liquid nitrogen but later during the study changed to a mixture of dry ice and isopenthane.

#### Exercise protocols

All participants exercised for 12 weeks, and intensity was based on results from the maximal exercise test (HRpeak). To monitor heart rate all participants wore a polar Unite watch and a chest heart-rate monitor (Polar H9) during all exercise sessions. Sessions were then synchronized to PolarFlow.com, to generic user accounts not related to the patient, where compliance as to frequency, intensity (HIIT group reaching >85 % and CON not surpassing 70 % of HRpeak) and duration of each session by the exercise instructor (HA).

#### Intervention group, HIIT

The HIIT protocol consisted of six interval sessions on a stationary bike, three times weekly, with each interval being 30-45 seconds long with two-minute-rest of moderate biking in-between. The heart rate was required to reach at least 85 % of HRpeak during each interval. Following the intervals, strength training of shoulders, knee extensors and core was performed on the load of 10 voluntary repetition maximum in one set. All strength training was progressive, when the rated exertion was lower than 7 on Borg CR-10, the resistance was increased if the participant could manage. Following one test session to identify correct initial exercise load/intensity, the training was ramped up during the first three to four weeks, starting with three sub-maximal intervals, then three maximal, and additional set were added on subsequently over three to four weeks until reaching the goal of six maximal intervals. Thereafter, the load or time of each interval was increased when needed to ensure that the heart rate exceeded 85 %. When the participant reached the goal intensity and was familiar with the protocol at least one session weekly needed to be supervised at the clinic and at least 75 % compliance was required. The participants were able to exercise at home or at a gym, if they wanted to, compliance and intensity being monitored by training journal and HR monitor.

#### Comparator group, low-moderate intensive home-based training, CON

This exercise protocol consisted of eight exercises of ten repetitions per exercise and external resistance could be added if necessary. It was performed five times weekly in combination with a 15-minute brisk walk where the participant could not surpass 70 % of HRpeak.

#### Mitochondrial analysis with Western Blot – Immunoblotting

Muscle tissues were homogenized in an appropriate volume of muscle extraction buffer: 50mM TrisCl pH 7.5, 150 mM NaCl, 1mM EDTA, 0.5 % NP40, 1 % SDS, 0.5 % DOC (fresh added: protease inhibitors, 1 mM DTT). Samples were centrifuged for 10 minutes, 12000g at 4°C. Clear supernatants were transferred in a new tube and protein content was measured using commercial Protein Assay kit (Bio-Rad, 5000001EDU). Approximatively 15 μg of protein for sample were incubated with an appropriate volume of loading buffer (Thermo-Fisher Scientific, NuPage LDS sample buffer NP0007, including denaturant agent B0009) and denaturate at 85°C for 5 minutes. The denaturation was not performed for samples used to detect the mitochondria respiratory chain proteins with anti-OxPhos antibodies. Samples were separated with NuPAGE SDS 4-12 % bis-tris gel and then protein transferred to PDVF membranes. Membranes were blocked with 5 % milk in TBS-0.1 % Tween20 for 1 h at room temperature, followed the overnight incubation with primary antibodies (rabbit anti-Citrate Synthase (1:1000) ab129095 AbCam, rabbit anti-VDAC1 (1:2000) ab15895 AbCam, and rodent anti-OxPhos (1:1000) 45-8099 Thermo Fisher Scientific) in cold room. All antibodies were diluted in blocking buffer. After, membranes were washed 5 times for 5 minutes each with TBS-0.1 % Tween20, then incubated with secondary antibody (1:5000, donkey anti-rabbit conjugated with HRP, DAKO) at room temperature for 1 hour. Membranes were again washed as before and HRP reagents were used to detect the bands using ChemiDoc system. Membranes were then stained with Coomassie blue to visualize the total protein loading. All images were analysed using ImageJ software.

### Procedure

The main author (KA), who was not involved in the treatment of patients, scheduled all appointments, supervised the maximal exercise test and was unaware of group allocation. The senior author (HA) was responsible for the exercise, both HIIT and CON, and led the sessions and instructed the participants. Blood tests were taken by the myositis team nurse (HS) who also randomized patients. A randomization table with blocks of four was used for randomizing participants to HIIT and CON, 1:1, and administrated by the objective myositis. A muscle biopsy was, when possible, taken by one rheumatologist, (MD, AN or IL). The same physician also performed the disease activity assessments before and after exercise and were blinded to group allocation. Patient demographics and characteristics were obtained through medical records or the Swedish Rheumatology Quality Registry (SRQ).

### Statistics and data analysis

Due to dropouts, there was missing data. This was handled in separate ways to perform two different analyses, intention-to-treat, and per-protocol. For the intention-to-treat analysis, all participants were included. Missing follow-up data from the maximal exercise test were replaced by the group mean change. Missing follow-up data for serum markers were assigned same value as at baseline, and for PhGA left as missing data for the statistical model to handle. For any missing baseline data, follow-up data were assigned missing as well. For the per-protocol analysis, only those with complete data and sufficient compliance were included. The results presented in the article are intention-to-treat, for per-protocol results and analysis, see Supplementary data S1 (sheet “Per-protocol data” and “Per-protocol CI”), and Supplementary data S3.

We analyzed the outcomes from the maximal exercise test, PhGA, and serum levels of muscle enzymes using R and Rstudio with a mixed linear model. This test was chosen as data was not normally distributed, the study is a repeated measures design, and the interaction of group*time is of essence. The model can handle skewness without transforming data, and missing data, while still giving robust output. Levels of mitochondrial protein expressions were analyzed within-group, using GraphPad and paired T-test. For statistical details, see Supplementary data S2.

## Results

### Exercise capacity

To compare the effectiveness between the two exercise interventions to improve aerobic fitness, we measured VO_2_peak at baseline and after 12 weeks. The change in VO_2_peak (L/min) was significantly larger for the HIIT group with a 16 % increase compared to 1.8 % for CON (95 % CI of 0.10;0.47; Fig. 2a; Table 2). Furthermore, VO_2_peak after normalization to bodyweight (mL/kg/min) was also significantly improved in the HIIT, 16 %, compared to the CON group, 4 % (95 % CI 0.83, 5.10), see Supplementary Figure S4; Table 2. When adjusting VO_2_peak for sex, weight, and age, most participants had a “very low” VO_2_peak at baseline, indicating a similarly impaired relative aerobic fitness among participants. A total of five participants improved their VO_2_peak in this measure (see Supplementary data S1, sheet “VO_2_ sex-age-weight-adjusted”).

**Table 2.** Results of aerobic capacity, muscle strength and disease activity assessments after 12 weeks of exercise.

| Variables | HIIT |  |  | CON |  |  | Between-group change interaction group*time, 95 % CI |
| --- | --- | --- | --- | --- | --- | --- | --- |
|  | Baseline (n=12) | 12-week follow-up (n=12) | Within-group change, 95 % CI | Baseline (n=11) | 12-w follow-up (n=11)* | Within-group change, 95 % CI |  |
| VO<sub>2</sub>peak L/min | 1.94 (1.59–2.17) | 2.22 (1.68–2.71) | **0.18;0.45** | 1.55 (1.3–1.86) | 1.5 (1.36–1.77) | -0.1;0.16 | **0.1;0.47** |
| VO<sub>2</sub>peak mL/kg/min | 24.8 (21.4–28.6) | 29.2 (25.4–32.5) | **2.34;5.49** | 22.1 (19.3–28.4) | 23.4 (19.8–29.8) | -0.6;2.5 | **0.83;5.1** |
| Peak power, W | 150 (120–166) | 188 (138–210) | **18;37** | 120 (92.5–148) | 130 (100–155) | **0.4;20.0** | **3.9;30.8** |
| Peak Heart rate | 171 (165 – 187) | 172 (162 – 187) | -6.5;2.0 | 157 (145 – 172) | 161 (149 – 169) | -4.0;4.0 | -7.9;3.7 |
| Time to exhaustion, mm:ss | 12:10 (09:12–13:38) | 15:59 (10:11–18:48) | **01:38;04:00** | 09:20 (06:50–11:16) | 10:27 (07:51–13:05) | -00:02;02:17 | **00:06;03:18** |
| MMT8, 0–80 | 80 (79 – 80)<sup>1</sup> | 80 (79.5 – 80) | -1.4;2.4 | 76.5 (73.5 – 80)<sup>1</sup> | 80 (78 – 80)<sup>1</sup> | **0.1;3.9** | -4.1;1.1 |
| Physician Global Activity, VAS 0–10 | 2.25 (1.5 – 3.0) | 1.2 (1.1 – 2.1)<sup>1</sup> | -1.30;0.04 | 2.0 (1.6 – 3.1)<sup>1</sup> | 0.9 (0.6 – 1.5)<sup>3</sup> | **-1.7;-0.26** | -0.6;1.2 |
| Muscle Disease Activity, VAS 0–10 | 0 (0.00–1.05) | 0.4 (0.00–1.25)<sup>1</sup> | -0.8;0.8 | 0.7 (0.00–0.95)<sup>1</sup> | 0 (0.0–0.3)<sup>3</sup> | -0.8;1.0 | -1.2;1.0 |

All data presented in median (interquartile range). All data analyzed with linear mixed model, within-group change for respective group is baseline vs. follow-up, between-group change is HIIT vs. CON. Significant results marked in bold. \*Three participants with missing follow-up data were assigned the group mean change as follow-up result. HIIT, high-intensity interval training; CON, control. 95 % CI, change is presented in 95 % confidence interval; VO<sub>2</sub>, peak oxygen uptake; L/min, liters per minute; mL/kg/min, milliliters per kilogram per minute; W, watts; time to exhaustion, time from start to finish; mm:ss, minutes:seconds; confidence interval for time to exhaustion in minutes:seconds; MMT8, manual muscle test 80; VAS, visual analog scale.

**Figure 2.**
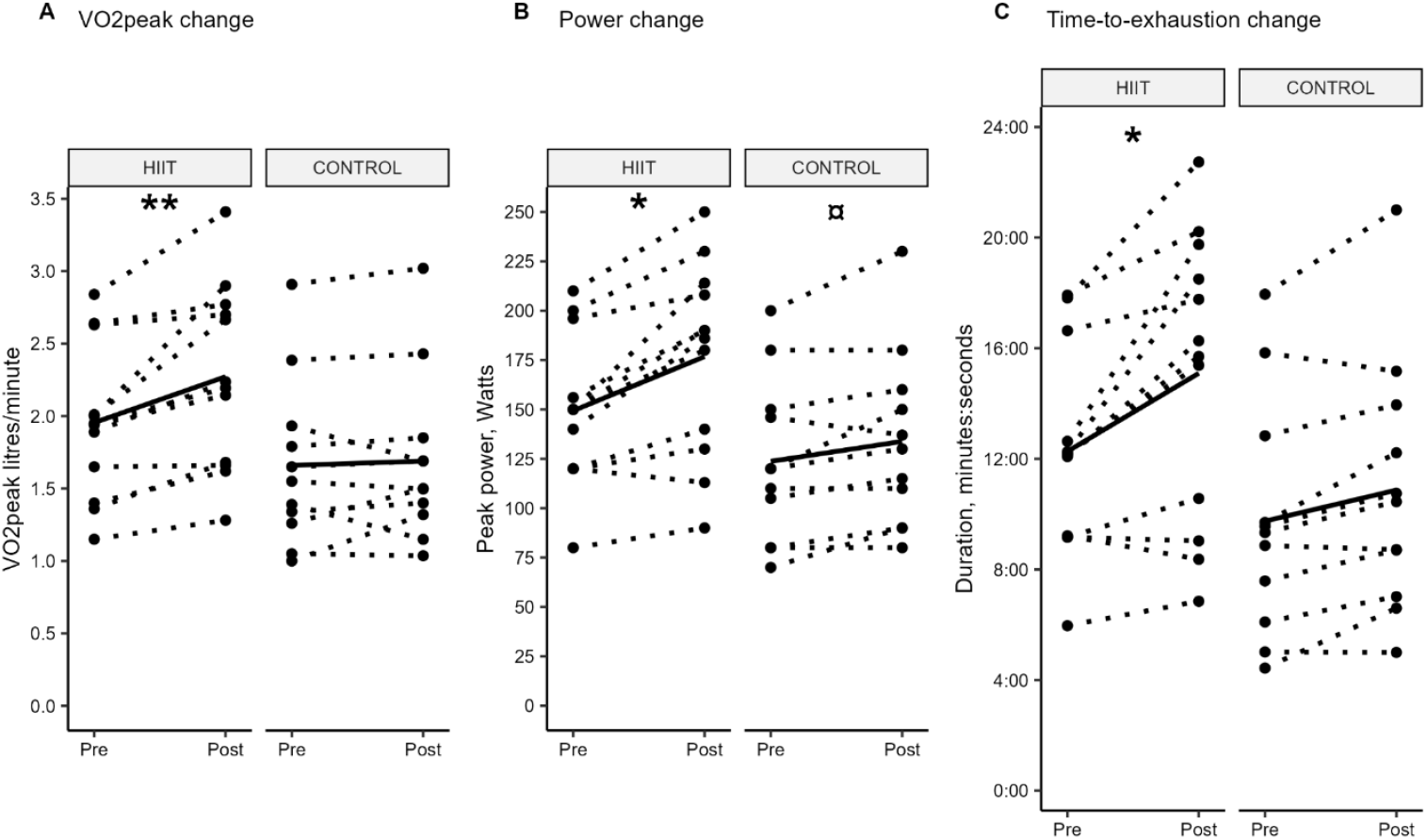
Results from maximal exercise test. Change from baseline (pre) and follow-up (post) per group. Dashed lines show change per participant, solid line shows group mean change. All data analyzed with linear mixed model, p-values are used for graphic simplicity, see 95 % CI in Table 2. VO_2_peak: peak oxygen consumption; HIIT: high-intensity interval training. * = between-group significance in favor of HIIT (*=p<0.05; **=p<0.01), ¤ = within-group significance.

Muscle endurance was improved in both peak power and TTE. Peak power was significantly improved in HIIT, 18 %, compared to CON, 8 % (95 % CI 3.9;30.8); however, the CON showed a significant within-group improvement as well (95 % CI 0.4;20.0), see Fig. 2b; Table 2. The HIIT group improved significant in time-to-exhaustion (TTE) during the maximal exercise test compared to the CON, HIIT 23 % compared to CON 11.25 % (95 % CI 6.0;198.0), see Fig. 2c; Table 2. Age-adjusted results remained significantly improved for VO_2_peak, and peak power remained in favor of HIIT, see Supplementary data S1 (sheet “Age adjusted data” and “Age adjusted CI”).

Heart rate at peak exercise (HRpeak) remained unchanged with respect to intervention in both groups (Table 2).

### Muscle mitochondrial adaptations

Aerobic exercise is a potent trigger of muscle adaptations including mitochondrial biogenesis. In conjunction to improved VO_2_max, we found the muscle expression of mitochondrial respiratory chain complexes I and V (CI & CV), citrate synthase (CS), and voltage-dependent anion channel 1 (VDAC1) increased in the HIIT (p<0.05), but not in CON. Protein quantification showed higher band intensities for these proteins post-intervention within HIIT but not within CON, indicating enhanced mitochondrial biogenesis and adaptations to aerobic metabolism (Figure 3).

**Figure 3.**
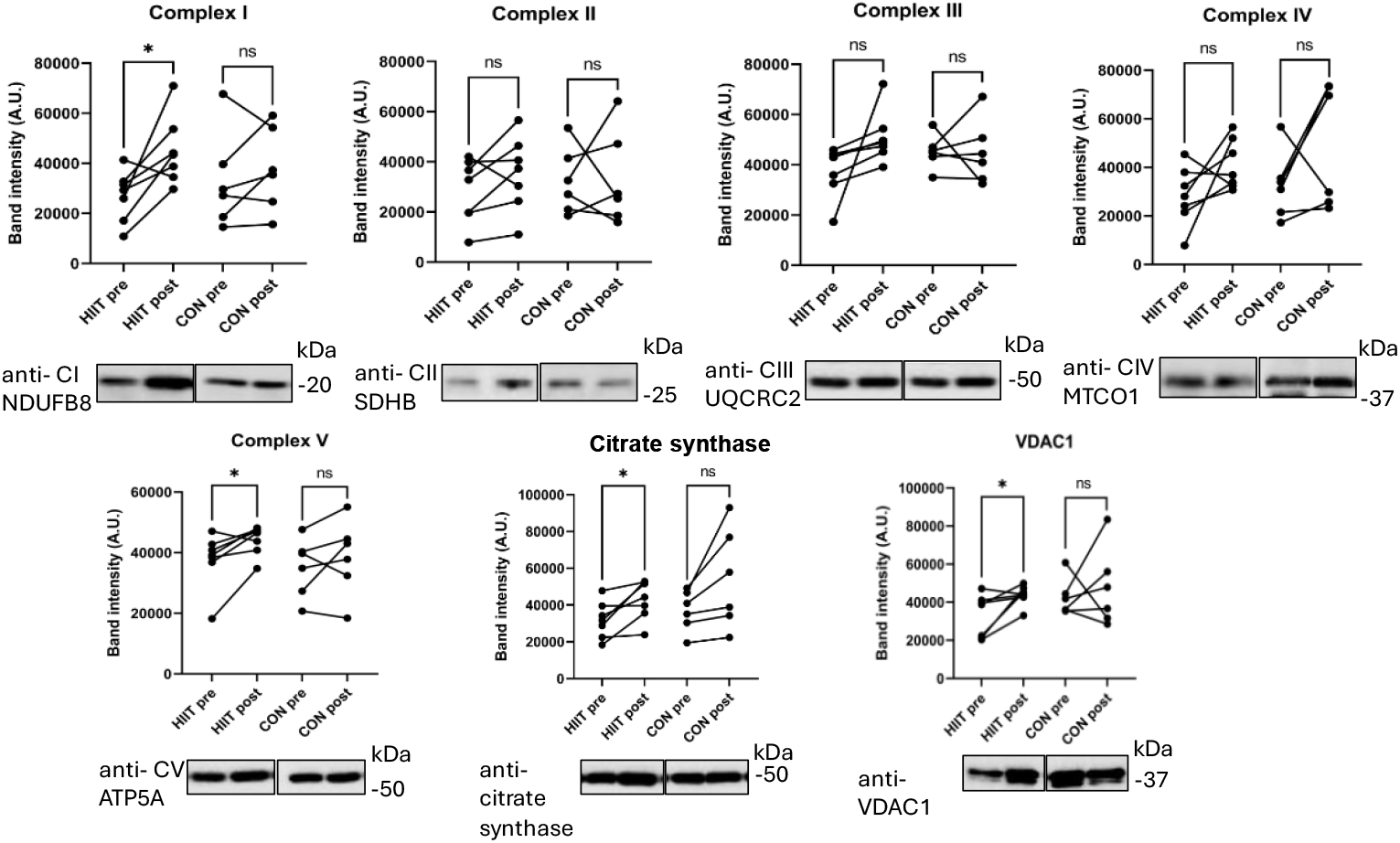
Increased performance after HIIT is accompanied with increased protein expression in the mitochondrial electron transport chain, citrate synthase (CS) and VDAC1 in skeletal muscle. Protein quantification for the mitochondria electron transport chain complexes I-V (A-E), citrate synthase (F), and VDAC1 (G) of skeletal muscle lysates from the High Intensity Interval Training (HIIT) group (n= 7) and the control (CON) group (n=6), before (PRE) and after (POST) (paired) exercises. Band intensity expressed in arbitrary units (A.U.) were determined by ImageJ and normalized to myosin as internal loading control. Representative blots and the specific antibodies to the quantified proteins: I-V (A-E) anti-OxPhos, anti-citrate synthase (F), and anti-VDAC1 (G) are indicated below the graphs. Paired t-test was performed PRE vs POST within the groups, * p<0.05. The uncropped blots are shown in the Supplementary Figure S5.

### Clinical evaluation scores

At baseline, the HIIT group already had the maximal Manual Muscle Test 80 (MMT8) score of 80 which was significantly better than the CON who displayed close to maximal scores (Table 2). The HIIT group remained at the maximal score after 12 weeks and the CON improved significantly (95 CI 0.11;3.89). PhGA significantly improved for CON. There was also a tendency towards improvement also in the HIIT group, although not statistically significant. MDAAT muscle disease activity remained unchanged in both groups (Table 2). There was no increase of serum levels of CK, AST or ALT after compared to before the HIIT or CON exercise. At baseline, two participants in HIIT had abnormally elevated CK levels; one remained equally elevated at follow-up, the other had normalized CK at follow-up. LD was slightly elevated in both groups at all time points; however, it did not increase after intervention (Table 1).

Out of the 23 participants, 20 completed the intervention. Two participants dropped out, and a third could not do follow-up tests due to COVID-19 restrictions, all allocated to the control group. There was a high compliance with a median 93 % of the possible HIIT sessions range 63-100 %. For the muscle biopsy analyses, a total of 13 participants were included with sufficient PRE and POST biopsy quality (HIIT n = 7, CON n = 6). Per-protocol analyses, compared to intention-to-treat revealed similar differences in favor of the HIIT except for a significantly higher HRpeak in HIIT compared to CON. For complete per-protocol analyses, age-adjusted results and weight-adjusted VO_2_peak, see Supplementary data S1.

## Discussion

In this first study to investigate efficacy and safety of HIIT compared to a standard-of-care exercise in recent onset IIM using a randomized controlled design, we found that HIIT was superior to current standard-of-care low-moderate exercise interventions to improve aerobic exercise capacity and muscle mitochondrial adaptations. Moreover, the results provide evidence that HIIT is safe and well tolerated in recent onset, IIM.

A notable finding was the significant increase in VO_2_peak in the HIIT group compared to the CON group. An increase >10 % in VO_2_peak is considered clinically relevant (34). In our study, the HIIT group improved VO_2_peak by 16 % corresponding to a clinically significant increase in VO_2_peak of 4.4 ml/kg/min. In contrast, the CON did not achieve statistically significant or clinically relevant change. Our results from the HIIT intervention are in line with effects of HIIT in the general population, where an average increase of 3.9 ml/kg/min was recently reported in a meta-analysis (35). The 4.4 ml/kg/min increase in the HIIT group is a substantial improvement exceeding 1 metabolic equivalent (MET) and underscores the efficacy of HIIT in enhancing aerobic capacity. This magnitude of increased VO_2_peak is known from other studies to associate with reduced all-cause mortality in healthy men and women (36).

Almost all participants in our study had a baseline VO_2_peak corresponding to “very low” aerobic capacity underlining the poor aerobic capacity in patients with IIM (34). Yet our HIIT protocol successfully improved aerobic capacity for all participants regardless of initial fitness level. To optimize adherence to the HIIT protocol also for unfit individuals, the initial exercise intensity was set low, based on the HRmax achieved at the baseline VO_2_ test, with gradual progression towards the goal intensity. Aerobic capacity is strongly correlated to self-reported physical health in IIM (37), which highlights the importance of assessing and targeting aerobic capacity in an exercise intervention for people with IIM. In the setting of cardiovascular diseases, improved aerobic capacity is strongly associated with reduced progression of cardiovascular disease and improved survival (38). To our knowledge it has not yet been studied how aerobic capacity links to disease outcome and mortality in IIM.

In addition to VO_2_peak, TTE and peak power also showed significant improvements in the HIIT group compared to the CON. HIIT-participants biked 23 % longer and achieved an 18 % increase in peak power, compared to 12 % and 8 % in the CON group, respectively. In this setting, we expected positive changes in peak power and TTE also in the CON as they also were exposed to exercise, although at a lower intensity. HRpeak did not change in either of the groups, suggesting that the participants reached near peak capacity at both baseline and follow-up.

Mitochondria is the central organelle for aerobic metabolism, electro transport chain, and many other metabolic processes in the muscle. Therefore, it is not surprising that mitochondrial dysfunction is linked to many diseases including IIM (39). It is well known that exercise training is a powerful signal for mitochondrial biogenesis (40) and, moreover, that VO2peak is associated to mitochondrial content in the muscle (41). In line with this, we found that repeated skeletal muscle biopsies revealed significant increases in mitochondrial protein expression following the HIIT intervention but not in CON. Increased level of expression was revealed in the electron transport chain (complex I and IV) proteins and for general markers of mitochondrial content (VDAC and citrate synthase). These proteins are crucial for mitochondrial function. The larger protein expression post-intervention in HIIT than CON, suggests that HIIT more effectively promotes mitochondrial adaptations in muscle, which may contribute to the observed improvements in aerobic capacity and muscle endurance. These molecular changes are consistent with previous research showing benefits of intensive exercise on mitochondrial function and overall metabolic health in healthy and in patients with IIM (21, 39, 42, 43).

We also assessed disease activity by measures of serum levels of muscle enzymes as surrogate markers of inflammation (CK, LD, AST, ALT), as well as the PhGA of disease activity and muscle disease activity. While CK levels were not elevated on a group level, LD was consistently elevated for both groups, suggesting ongoing muscle damage or stress. However, LD remained unchanged, suggesting no increased inflammation from either HIIT or CON supporting the safety of HIIT.

Interestingly, the CON group showed a significant improvement in PhGA. Further, CON also had a slight room for improvement in MMT scores which HIIT did not as they had maximal scores already at baseline. Muscle disease activity was scored slightly higher, although non-significant, for the HIIT group at follow-up. However, the change from 0 cm to 0.7 cm is within the error of measurement and probably not clinically relevant. Therefore, our results are consistent with previous studies on exercise in IIM (24); as there were no signs of increased inflammation or disease activity, suggesting HIIT being a safe intervention also in early phases of disease.

The HIIT group had a maximum score of 80 on MMT8 and thus there was no room for improvement. The CON had almost 96 % of maximal score at baseline and improved significantly, after 12 weeks. Overall, the clinical relevance or importance of MMT8, with its previously described ceiling effects (4, 44), can be discussed as it might lead to overseen limitations in muscle function in patients. Unfortunately, we were not able to perform FI-2, as it would have added another separate visit to the clinic. However, the results of MMT8 indicate no worsening of muscle strength further underscoring the safety of HIIT.

Despite the rigorous and intensive nature of HIIT, compliance was high, and the protocol was well-tolerated by participants. This is encouraging, as it demonstrates the feasibility to implement HIIT in a clinical setting for IIM patients. The use of heart rate monitors and careful supervision ensured that participants adhered to the prescribed intensity levels, minimizing the risk of adverse events.

The study had some limitations, including missing data for some participants. However, the use of intention-to-treat and per-protocol analyses helped mitigate these limitations. Furthermore, the blinded randomization and use of control group strengthens our results. Despite randomization in blocks of four there was a skewness, although non-significant, in age, sex, and type of IIM between the two groups (Table 1).

The study was initiated as a pilot study since this type of intensive exercise had never been evaluated in patients with recent onset IIM. COVID-19 restrictions forced us to change the inclusion criteria of time from diagnosis from 6 months to 12, to be able to include four patients that were about to start exercise at the time of initiating pandemic restrictions. Further, the pandemic hindered us from including at least two patients with higher disease activity and organ involvement as they were “high-risk patients” and were not allowed into the hospital other than for clinically necessary appointments. In this study we were not able to retrieve paired biopsies from all participants. Initially, this was due to staff shortage and a large re-organization to a newly built hospital with lack of routines. Later, it was also due to freezer breakdowns leading to thawed biopsies, and in some cases, the biopsies consisted also of fat with too little muscle tissue for the analyses.

## Conclusions

This study demonstrates that HIIT is a superior exercise regimen compared to standard low-moderate intensity home-based exercise for improving aerobic capacity, muscle endurance, and mitochondrial adaptation in patients with IIM with short disease duration. These findings support the use of HIIT as a safe and effective adjuvant treatment in the management of IIM, which provides an effective strategy to enhance physical fitness and potentially improve quality of life for these patients. Future studies should aim to explore long-term effects of HIIT on both physical fitness, disease activity and possibly the role of HIIT to prevent comorbidities and mortality. Additionally, ongoing investigation of underlying mechanisms of HIIT’s impact on inflammation and mitochondrial function could provide valuable insights into the pathophysiology of IIM.

## Data Availability

All data produced in the present study are available upon reasonable request to the authors

## Notes

### Competing Interest Statement

I.E.L. has received research grants from Astra Zeneca and Janssen Pharmaceutica NV and has been serving on the advisory board for EMD Serono Research & Development Institute, Argenx, Pfizer, Galapagos, Chugai Pharmaceutical Co., Ltd; Novartis, Bristol Myers Squibb and Janssen Pharmaceutica NV and has stock shares in Roche and Novartis.

### Clinical Trial

NCT03324152

### Funding Statement

I.E.L was supported by Swedish Research Council (2020-01378), ALF (FoUI-955086), Heart and Lung foundation (20220127) King Gustaf V 80-year foundation and Swedish Rheumatism Association.
D.C.A was supported by King Gustaf V 80-year foundation (FAI-2022-0935),
Stiftelsen Promobilia (A23059), Stockholm County research grant (ALF;
FoUI-1002205) and Swedish Heart-Lung Foundation (20230594).
H.A was supported by Swedish Research Council, Swedish Rheumatism Association, Promobilia Foundation, ALF (Agreement between central government and Stockholm Region)

### Author Declarations

The Regional Ethics Review Board in Stockholm gave ethical approval for this work

