## Supplementary material for "High-Intensity Interval Training Outperforms Moderate Exercise to Improve Aerobic Capacity in Patients with Recent-Onset Idiopathic Inflammatory Myopathies: A Multicenter Randomized Controlled Trial": S2 - supplementary data 2

### Statistical analysis in RStudio

**Packages used in analysis and plotting:**
lme4, Matrix, lmerTest, ggplot2, dplyr, writexl, hms, tidyr, emmeans, ggpubr, patchwork, rstatix, ggbreak

**Code for Mixed Linear Models:**
vo2L <- lmer(vo2.L ~ Randomisation * Timepoint + (1 | ID), data = dfhiit.itt)

- “vo2L” is the name of the parameter that will contain the output of the formula
- “vo2.L” is the variable analyzed
- “Randomisation” & “Timepoint” are fixed effects
- * creates an interaction between “Randomisation” & “Timepoint”
- “(1 | ID)” is the random intercept, it allows for baseline values of vo2.L to vary between subjects
- “data = dfhiit.itt” is the dataset containing all variables in the program

**Code for age adjusted Mixed Linear Models:**

Vo2L.age <- lmer(vo2.L ~ Randomisation * Timepoint + age + (1 | ID), data = dfhiit.itt.age)

- All similar as above but the variable “age” is added as a fixed effect (covariate), meaning the model assumes age will have an effect on VO2.L
