## Supplementary material for "High-Intensity Interval Training Outperforms Moderate Exercise to Improve Aerobic Capacity in Patients with Recent-Onset Idiopathic Inflammatory Myopathies: A Multicenter Randomized Controlled Trial": S3 - supplementary data 3

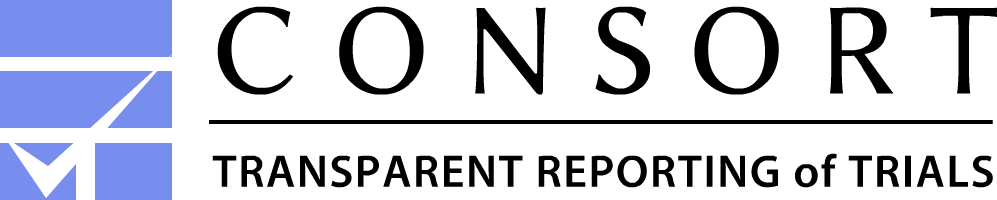

**CONSORT 2010 Flow Diagram**

**Allocation**

**Analysis**

**Follow-Up**

**Enrollment**

Assessed for eligibility (n=82)

Excluded (n=59)

♦  Not meeting inclusion criteria (n=24)

♦  Declined to participate (n=15)

♦  Other reasons (n=20)

Analysed (n=11)
♦ Excluded from analysis (n=1)
 •Insufficient compliance (n=1)

Lost to follow-up (give reasons) (n=0)

Discontinued intervention (n=0)

Allocated to intervention (n=12)

♦ Received allocated intervention (n=12)

♦ Did not receive allocated intervention (n=0)

Lost to follow-up (n=1)

♦ Covid-19 restrictions (n=1)

Discontinued control (n=2)
♦ Excessive exercise (n=2)

Allocated to control (n=11)

♦ Received allocated control (n=11)

♦ Did not receive allocated control (n=0)

Analysed (n=8)
♦ Excluded from analysis (n=3)
Missing data (n=0)
♦ No follow-up/discontinued control (n=3)

Randomized (n=23)
