## Supplementary figures and images for "High-Intensity Interval Training Outperforms Moderate Exercise to Improve Aerobic Capacity in Patients with Recent-Onset Idiopathic Inflammatory Myopathies: A Multicenter Randomized Controlled Trial"

### S4 - supplementary figure 1

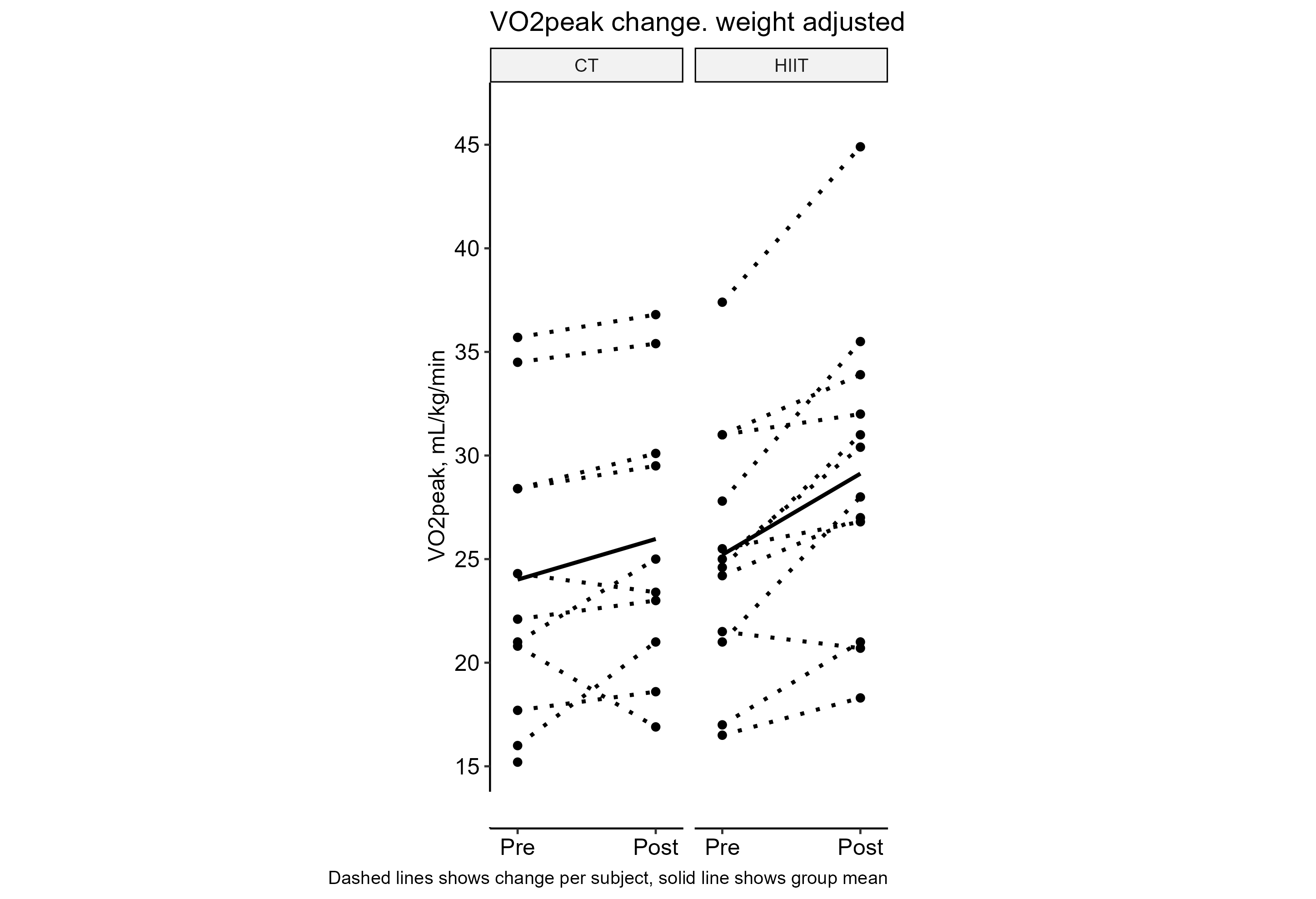
