## Supplementary material for "High-Intensity Interval Training Outperforms Moderate Exercise to Improve Aerobic Capacity in Patients with Recent-Onset Idiopathic Inflammatory Myopathies: A Multicenter Randomized Controlled Trial": S5 - supplementary figure 2

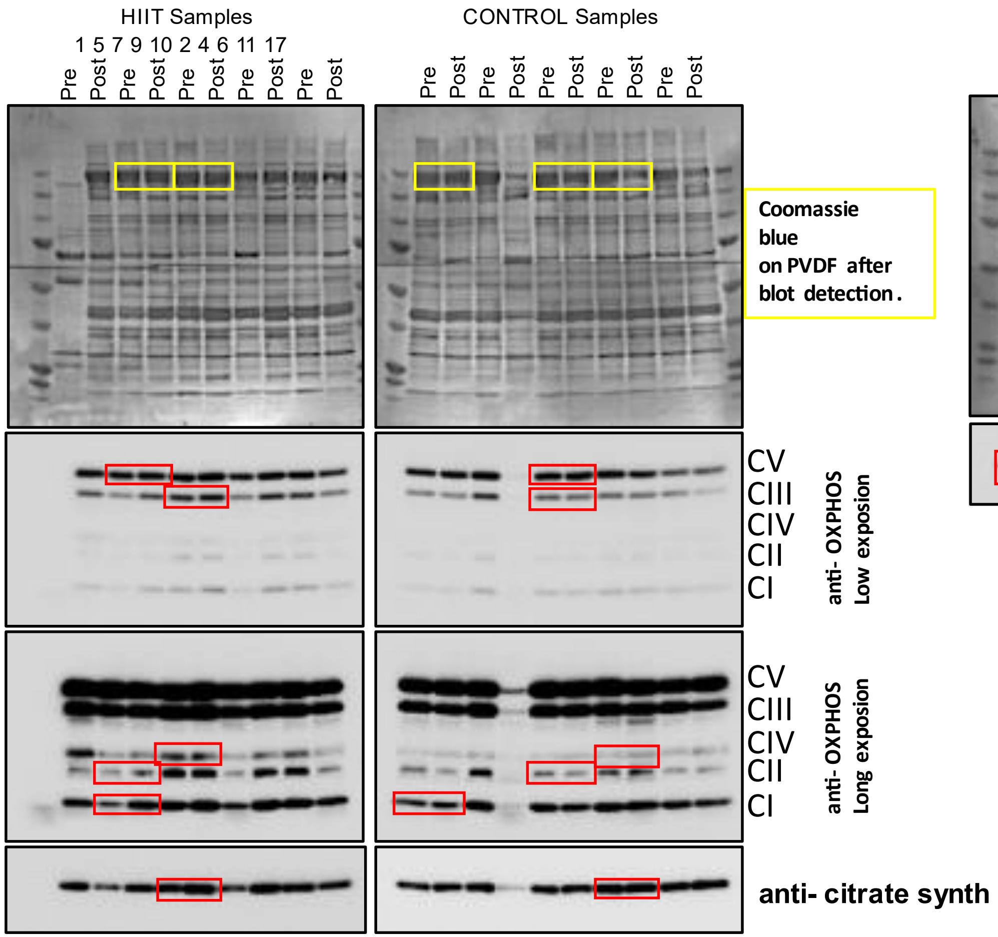


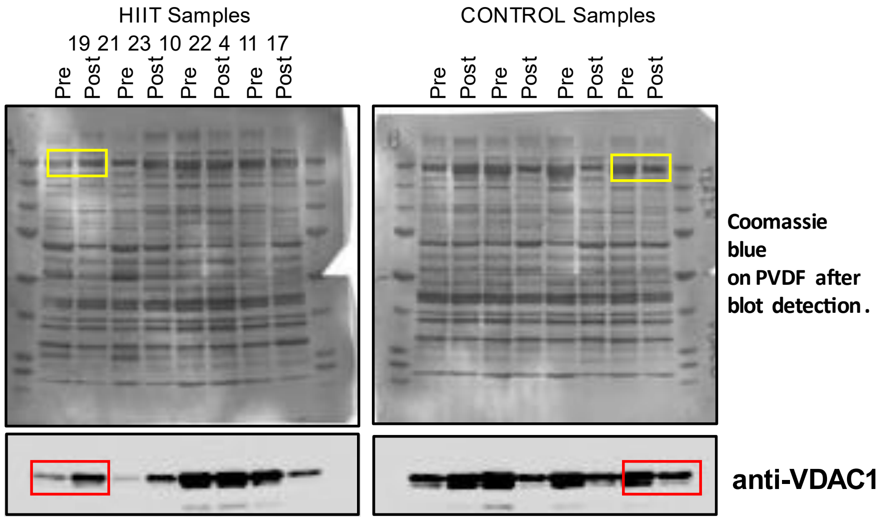


15 µg of total muscle proteins of each samples were loaded per well. Wester blots and coomassie blue on PVDFmembranes after blot detection.

In red are indicated the cropped areas showed in the Figure X and in yellow the myosin band used for loading normalization.
